# Genomic analysis identifies polygenic and region-specific contributions to ADHD-migraine comorbidity

**DOI:** 10.64898/2026.08.19.26360780

**Authors:** Yaxin Luo, Christina Dardani, Robyn E Wootton, Evie Stergiakouli

## Abstract

Attention-deficit/hyperactivity disorder (ADHD) and migraine frequently co-occur, yet their shared genetic architecture remains unclear. Using genome-wide association summary data of ADHD and migraine, we applied a multi-layered genetic analysis framework. Conjunctional false discovery rate was used to identify shared pleiotropic variants. Local genetic correlation was performed within semi-independent regions. Mendelian randomization (MR) used eQTLs as instruments to assess overlap of genetically predicted gene expression on ADHD and migraine in relevant brain and blood tissues. Colocalization analyses were conducted to assess whether shared association signals were driven by the same underlying variants. We identified 27 pleiotropic variants shared between ADHD and migraine, 20 with concordant effect directions. Local genetic correlation analysis identified a single shared region on chromosome 11 with evidence of local heritability for both ADHD and migraine and a positive local genetic correlation. Cis-eQTL MR of druggable genes identified multiple genes with evidence for causal effects of their expression on both ADHD and migraine across brain cortex and blood. Genetically predicted *MANBA* expression showed consistent associations with both traits in brain cortex and blood. Colocalization for *MANBA* supported a shared causal variant in cortex but not in blood, suggesting tissue-specific mechanisms. Current findings provide evidence for shared genetic architecture between ADHD and migraine across variant, regional, and gene-expression levels. Among our findings, genetically predicted expression of *MANBA* in the brain cortex appeared to be a potential shared biological contributor to ADHD and migraine.

## Introduction

Attention-deficit/hyperactivity disorder (ADHD) is a neurodevelopmental condition frequently accompanied by a wide range of psychiatric and somatic comorbidities that can affect functioning and quality of life [1, 2]. Epidemiological studies have consistently reported an association between ADHD and migraine, with several studies demonstrating a higher prevalence of migraine among individuals with ADHD than among those without ADHD [3, 4]. This association appears to be specific to migraine rather than headache disorders in general: a recent meta-analysis including cohorts from Europe and the United States reported a modest but migraine-specific association, with no corresponding evidence of association for tension-type or general headache disorders [5]. Similar associations have also been reported in independent community samples. For example, in the Danish Blood Donor Study, self-reported migraine was associated with a higher likelihood of ADHD, with the strongest associations observed among individuals reporting visual aura [6]. Evidence from intergenerational designs also pointed in the same direction: a population-based registry data in Taiwan found that parental migraine was associated with elevated ADHD risk in offspring [7]. Together, these findings across multiple designs and populations suggest a consistent pattern of ADHD-migraine co-occurrence, although observational data alone cannot clarify the underlying mechanisms.

Both ADHD and migraine are moderately to highly heritable conditions. Twin and family studies indicate that ADHD is highly heritable, with heritability estimates typically ranging from 70% to 80% [8]. Migraine also aggregates strongly within families, with risk particularly elevated among first-degree relatives and in families with multiple affected members [9]. Genetic studies also provide an additional line of evidence supporting a link between the two conditions. Large cross-trait genomic analyses have reported a significant genetic correlation between the two conditions (*r_g_*□=□0.26, *P*□=□8.81□×□10^−8^), suggesting that part of their inherited risk is shared [10]. The same analyses also identified genetic overlap between ADHD and several cognitive and psychiatric traits, including educational attainment (EA) and major depressive disorder (MDD), indicating that broader cognitive and emotional factors may contribute to this relationship. Complementary findings from a Swedish population-based cohort show that individuals with higher polygenic risk for ADHD have a modestly increased likelihood of developing migraine later in life (OR = 1.07, 95% CI = 1.02–1.12) [11]. Prior work has also explored variant-level pleiotropy between the two conditions by examining cross-trait genetic overlap and shared risk variants across ADHD and migraine. These studies reported evidence of shared association signals, suggesting partial overlap in genetic architecture. However, these analyses relied on earlier and substantially smaller ADHD GWAS datasets and therefore provided only an initial characterization of shared genetic architecture [12].

Despite these advances, a critical gap persists in understanding genomic and regulatory mechanisms underlying this comorbidity. The specific genetic loci underpinning this shared heritability remain largely unknown, and it is unclear whether observed cross-trait correlations reflect shared causal variants, distinct variants in linkage disequilibrium (LD), or pleiotropic effects on related biological processes. While prior candidate gene studies have implicated overlapping pathways in neurotransmission (e.g., dopaminergic and serotonergic systems involving *DRD4*, *DRD5*, *DAT1*, and *HTR1B*) [13, 14], such approaches were limited in scope in that they focused on a small number of *a priori* selected genes and therefore could not capture the highly polygenic nature of the shared genetic architecture. Moreover, no study has systematically integrated variant-level, regional, and tissue-specific cis-eQTL analyses to characterize whether shared genetic signals converge on biologically or therapeutically relevant pathways.

To better characterize the genetic overlap between ADHD and migraine, we combined variant-level, regional-level, and gene-level analyses to systematically investigate shared genetic architecture between ADHD and migraine. This integrative strategy enables a multilayered evaluation of the genetic mechanisms linking the two conditions.

## Materials and Methods

### Study Design Overview

To systematically investigate the genetic overlap between ADHD and migraine, we applied three analytical approaches that address complementary but distinct questions (Figure 1): (1) Conjunctional false discovery rate (ConjFDR) to identify pleiotropic SNPs that associate with both migraine and ADHD; (2) Local analysis of variant association (LAVA) to assess whether shared genetic signal between ADHD and migraine is concentrated within specific LD-defined genomic intervals; and (3) Mendelian Randomization (MR) analysis at the gene level to evaluate whether genetically predicted expression of the same genes with potential therapeutic relevance causally influence both ADHD and migraine.

**Figure 1.**
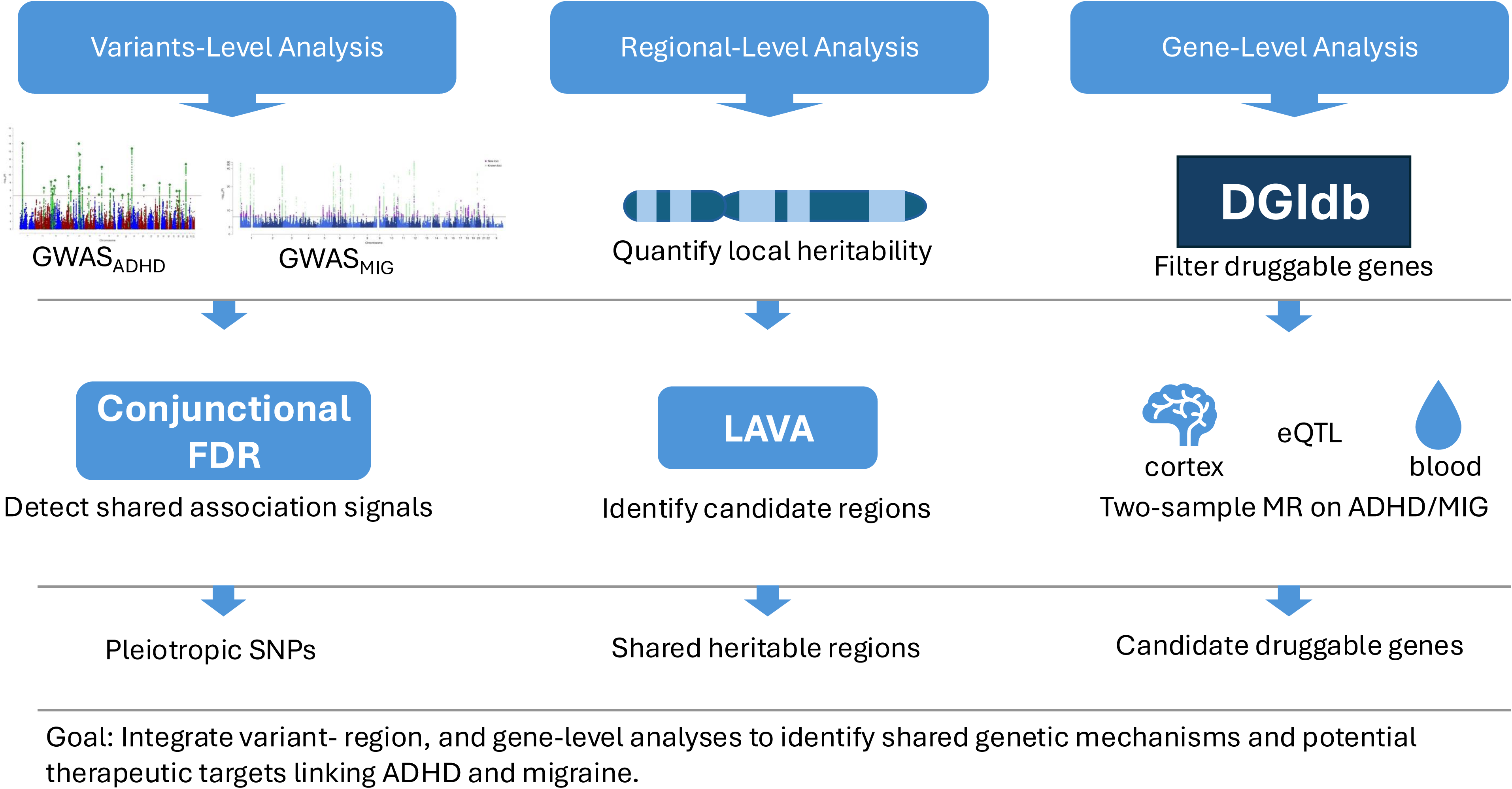
Analytical workflow for variant-, regional-, and gene-level analyses of ADHD and migraine. All analyses started from the same quality-controlled and harmonised ADHD and migraine GWAS summary statistics.

### Data Sources and Quality Control

We utilized summary statistics from the largest publicly available GWAS for each phenotype. Migraine GWAS summary statistics were obtained from a meta-analysis of 48,975 cases and 540,381 controls, integrating four cohorts (IHGC 2016, UK Biobank, GeneRISK, and HUNT) [15]. ADHD GWAS summary data were sourced from the latest meta-analysis (N = 296,487), which combined GWAS results of quantitative ADHD symptom scores and clinical ADHD diagnosis [16]. Quantitative ADHD symptom scores derived from multiple validated instruments, primarily ASEBA- and the Strength and Difficulties Questionnaire (SDQ)-based assessment. For diagnosed ADHD, cases are defined as clinically diagnosed with ADHD or prescribed mediation specific to ADHD. More details on the data sources are presented in Table S1.

Quality control and harmonization was performed using the 1000 Genomes Phase 3 European ancestry reference panel [17]. We retained SNPs with minor allele frequency ≥ 1% in both the reference and GWAS datasets. Ambiguous SNPs with allele frequency differences > 0.2 were excluded. To mitigate confounding effects due to the long-range linkage disequilibrium and complex genetic architecture of the major histocompatibility complex (MHC) region, we excluded all variants within the extended MHC region on chromosome 6 (chr6: 25,119,000 - 33,855,000, GRCh37/hg19) from the summary statistics. After harmonization of alleles and strand orientation, 6,302,141 autosomal SNPs common to both traits were retained for subsequent analyses. All subsequent analyses were conducted on this harmonized and MHC-excluded SNP set.

#### Analysis 1: Variant-Level Analysis

Conditional Q-Q plot were used to assess cross-trait enrichment by examining whether SNPs increasingly associated with ADHD showed stronger enrichment for migraine associations, and vice versa. We applied the conjFDR method using overlapping ADHD and migraine GWAS summary statistics to identify SNPs jointly associated with both traits [18]. ConjFDR quantifies cross-trait enrichment by estimating conditional FDR in both direction, conditioning ADHD associations on the strength of migraine associations and vice versa. SNPs were considered jointly associated with ADHD and migraine when the conjunctional FDR, defined as the maximum of the two conditional FDR values, fell below the predefined threshold.

Variants with conjFDR < 0.05 were considered significantly pleiotropic. As conjFDR identifies pleiotropic association signals without resolving whether they arise from a shared causal variant or distinct variants in LD, we performed pairwise conditional colocalization (PWCOCO) to test whether ADHD and migraine share a causal variant [19]. In this framework, posterior probability for H4 (PP.H4) represents the hypothesis that the same underlying causal variant drives the association signals for both traits at a given locus, rather than two distinct but nearby variants. We considered variants with H4 > 0.8 as providing strong evidence of colocalization, and those with PP.H4 > 0.7 as providing suggestive evidence. To aid interpretation of colocalized variants (PP.H4 > 0.8), we performed variant-to-gene annotation using MAGMA. MAGMA aggregates SNP-level association statistics into gene-level tests, providing a statistical summary of whether genes in the region show aggregated evidence of association, thereby highlighting candidate genes potentially underlying the shared genetic architecture [20] (Implementation details are provided in Supplementary Methods S1).

#### Analysis 2: Region-Level Analysis

Because both traits are highly polygenic, shared genetic architecture may also be reflected at the regional level rather than solely at individual variants. We therefore applied LAVA (v0.1.5) [21] to estimate local heritability and local genetic correlation between the two traits across predefined LD blocks. This framework enables the identification of genomic regions contributing to local genetic overlap between the two traits.

Analyses were performed across approximately 2,495 semi-independent LD blocks defined using the default genome partitioning scheme provided by LAVA [21]. Local heritability and local genetic correlation were estimated using the UK Biobank European reference panel [22]. For each LD-block, we first estimated local heritability for ADHD and for migraine. Regions showing evidence of heritability for both traits were then tested for local genetic correlation, with significance assessed using Bonferroni correction (P<0.05/2495). Given that EA and MDD are genetically correlated with both ADHD and migraine [10], we additionally performed conditional local genetic correlation analyses adjusting for EA [23] and MDD [24]. For all LAVA analyses, the cross-trait intercept was estimated from LD score regression and was used as the sampling covariance input [25], in order to account for correlation between GWAS summary statistics arising from sample overlap or other shared sources of confounding.

To summarize association patterns within LAVA-identified regions, we performed a region-restricted gene-based analysis using MAGMA. This approach prioritizes genes located within LD-defined regions showing significant local genetic correlation between ADHD and migraine (Implementation details are provided in Supplementary Methods S2).

#### Analysis 3: Gene-Level Analysis

We defined pharmacologically actionable (“druggable”) genes using the Drug-Gene Interaction Database (DGIdb v5.0), which aggregates information on known drug targets and potentially druggable genes from multiple curated resources [26]. This provides a comprehensive catalogue of genes that are known to be or potentially are targetable by existing compounds.

We utilized cis-eQTL data from two large-scale consortiums: brain cortex (2,683 samples from the MetaBrain consortium) [27] and whole blood (31,684 samples from the eQTLGen consortium) [28], both comprising primarily individuals of European ancestry. The cortex was selected as the representative brain tissue because of its established involvement in cognitive and pain-related processes implicated in ADHD and migraine [29, 30]. For each druggable gene, we first identified significant cis-eQTLs located within a 1 Mb window upstream of the transcription start site and downstream of the transcription end site. Independent instruments were obtained by LD clumping (r² < 0.001) among genome-wide significant cis-eQTL (P < 5×10□□), ensuring that only independent cis-acting variants were used in the MR analysis. For IVs that were not present in the outcome GWAS, proxies were identified as r2>0.8 with nearest physical distance using the ‘LDlinkR’ package with the 1000 Genomes European reference panel.

For each tissue (brain and blood), we performed two separate MR analyses, testing whether genetically predicted gene expression influenced ADHD and migraine. Genetically predicted expression of a gene was considered to show overlapping evidence across traits within a given tissue if it passed FDR correction for both trait (e.g. brain to ADHD and brain to migraine). The same criterion was applied independently for blood.

To distinguish shared causal variants from LD, we applied PWCOCO to test whether the cis-eQTL signal and the trait association were driven by the same causal variant. As before, PP.H4 > 0.8 was interpreted as strong evidence of colocalization, and PP.H4 > 0.7 as suggestive evidence (Implementation details are provided in Supplementary Methods S3).

## Results

### Analysis 1: Variant-Level Analysis

Conditional Q-Q plots revealed substantial upward deviations in the tail of the distribution, supporting the presence of shared genetic architecture between the two traits (Figure S1). This pattern was bidirectional, observed when conditioning ADHD on migraine and vice versa, providing initial evidence of widespread polygenic overlap.

Applying a conjunctional FDR threshold (conjFDR < 0.05) and pruning for linkage disequilibrium (LD; r² > 0.2), we identified 27 independent pleiotropic variants between ADHD and migraine (Table S2, Figure 2) and 20 of them showed consistent direction of effects across both phenotypes. Pairwise conditional colocalization analysis revealed that among those 20 variants, two showed strong evidence of colocalization, rs56262138 (H4 = 0.94) and rs2431108 (H4 = 0.87, Figure S2-3) and one variant showed suggestive evidence (rs12896360, H4 = 0.71). The full results of colocalization analyses are presented in Table S3.

**Figure 2.**
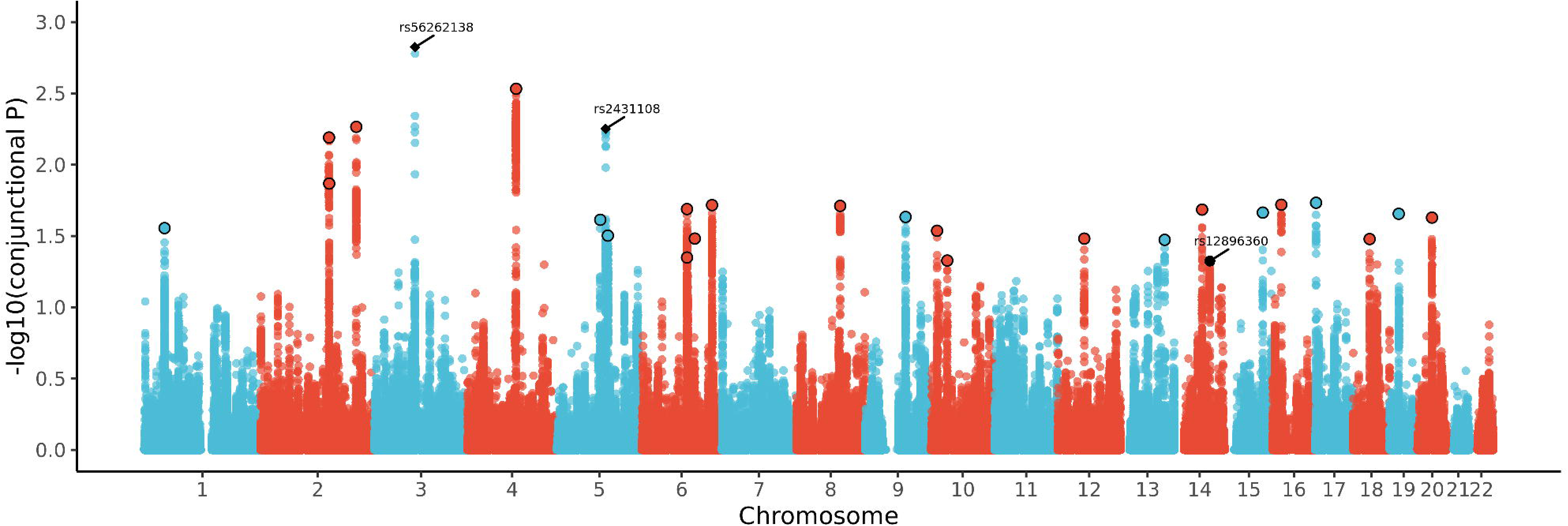
Common genetic variants jointly associated with ADHD and migraine at conjunctional false discovery rate (conjFDR)<0.05. Manhattan plot showing the - log_10_ transformed conjFDR values for each SNP on the y-axis and chromosomal positions along the x-axis. Independent lead SNPs are outlined in black. Filled circles indicate suggestive colocalization (0.7 < PP.H4 ≤ 0.8), whereas filled diamonds indicate strong colocalization (PP.H4 > 0.8).

To prioritize candidate genes in regions showing evidence for shared causal variants (H4>0.8), we performed MAGMA gene-based association analysis within 1Mb of each SNP. Two genes *CADM2* and *VGLL3* were tagged by rs56262138. *CADM2* showed gene-level association signals for both ADHD (P = 3.05 × 10□^8^) and migraine (P = 4.95 × 10□^4^), whereas *VGLL3* was primarily associated with migraine (P = 9.12 × 10□^4^) and exhibited a weaker signal in ADHD (P = 5.05 × 10□^3^, Table S4). No gene was annotated to variant rs2431108.

### Analysis 2: Regional-Level Analysis

We next assessed local genetic correlations between ADHD and migraine using the LAVA framework, which partitions the genome into approximately independent LD blocks and tests for shared genetic effects at the regional level.

At a Bonferroni-corrected threshold (P<0.05/2495), evidence for univariate local heritability was detected in 53 regions for ADHD and 21 regions for migraine (Table S5). Among these, only one genomic region, located on chromosome 11 (chr11:112755447-113889019), showed evidence for local heritability for both traits and was therefore taken forward for bivariate analysis. In this region, we observed a positive local genetic correlation between ADHD and migraine (rho = 0.53, 95% CI = 0.24, 0.88, P = 8.59× 10□□), indicating shared genetic influence at this locus. After conditioning on EA, the estimate became unstable and imprecise, with the confidence interval reaching the parameter boundary (partial ρ = 1.00, 95% CI: 0.11–1.00; P = 0.058). Adjustment for MDD retained nominal evidence of local genetic correlation, although precision was reduced (partial ρ = 0.55, 95% CI = 0.04, 1.00; P = 0.036). When jointly adjusting for both EA and MDD, the local genetic correlation was further attenuated (partial ρ = 1.00, 95% CI = −0.44, 1.00; P = 0.12, Table S6).

To further characterise the shared regional signal, we performed gene-based association analyses within this interval using MAGMA (Table S7). Several genes within this region showed stronger gene-level associations with ADHD than with migraine, including *USP28*, *NCAM1*, and *ZW10*, whereas corresponding associations with migraine were generally weaker and did not reach comparable levels of evidence. To visualise the underlying SNP-level association structure with the same interval, we plotted regional GWAS association profiles for ADHD and migraine (Figure 3). This profile shows that ADHD exhibits stronger SNP-level signals across the region, while migraine displays more modest associations.

**Figure 3.**
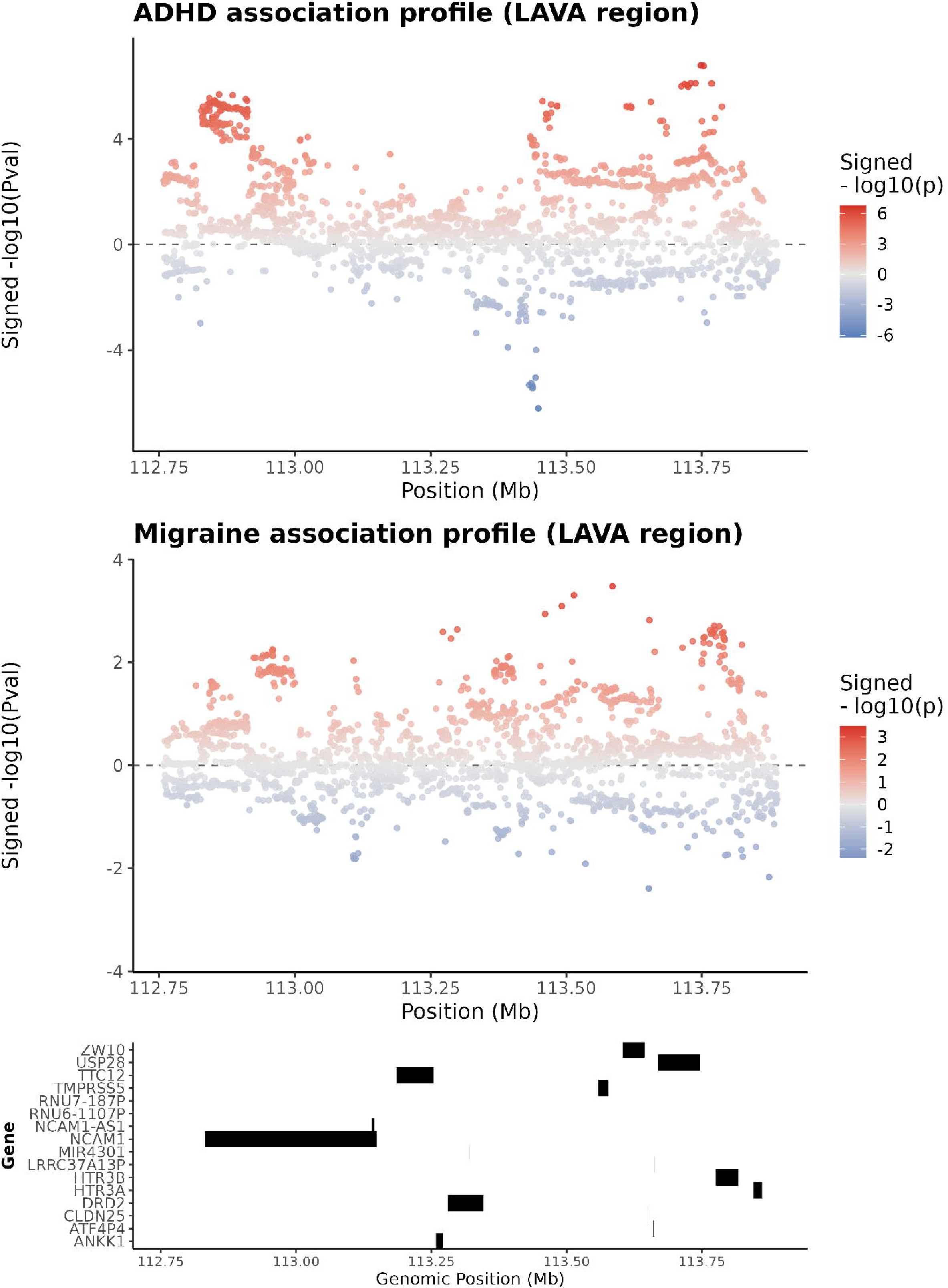
Regional association profiles for ADHD and migraine within the shared chromosome 11 region exhibiting significant local genetic correlation. SNPs are coloured according to the direction and strength of association, with red indicating positive effect estimates and blue indicating negative effect estimates.

### Analysis 3: Gene-Level Analyses

Following quality control, instrument sets were available for genetically predicted expression of 2,195 brain-derived and 2,116 blood-derived druggable genes for subsequent MR analysis (Table S8-9). Using cis-eQTLs from the MetaBrain (cortex) dataset, we found 34 genes whose genetically predicted expression showed MR evidence consistent with a potential causal effect on ADHD and 8 for migraine (Table S10-S11). Analyses based on blood cis-eQTLs from the eQTLGen consortium yielded 15 genes whose genetically predicted expression showed MR evidence consistent with a potential causal effect on ADHD and 6 for migraine (Table S12-S13).

Among the genes identified in brain cortex-based analyses as showing evidence of association with either ADHD or migraine, genetically predicted *MANBA* expression in cortex showed evidence for both ADHD and migraine (ADHD: OR = 0.94, 95% CI = 0.92, 0.97; Migraine: OR = 0.87, 95% CI = 0.81, 0.93). In blood-based analyses, *MANBA* also showed evidence of association with both traits (ADHD: OR = 0.95, 95% CI = 0.92, 0.97; migraine: OR = 0.88, 95% CI = 0.83, 0.94; Table 1).

**Table 1.** Genetically predicted expression of MANBA with evidence of causal effects on both ADHD and migraine based on cis-eQTL MR analyses in brain and blood tissues.

| Tissue | Outcome | OR (95% CI) | P value | H3 | H4 |
| --- | --- | --- | --- | --- | --- |
| Cortex | ADHD | 0.94 (0.92, 0.97) | $2.92 \times 10^{-6}$ | 0.23 | 0.77 |
| Cortex | Migraine | 0.87 (0.81, 0.93) | $2.24 \times 10^{-5}$ | 0.20 | 0.73 |
| Blood | ADHD | 0.95 (0.92, 0.97) | $1.31 \times 10^{-6}$ | 0.98 | 0.001 |
| Blood | migraine | 0.88 (0.83, 0.94) | $3.13 \times 10^{-5}$ | 0.77 | <0.001 |

To assess whether these MR relationships were likely driven by a shared causal variant, we performed pairwise colocalization between *MANBA* expression and each trait from cortex and blood, separately. In cortex, PWCOCO indicated suggestive evidence for a shared causal variant in cortex (*MANBA* expression in cortex with ADHD: H□ = 0.77; with migraine: H□ = 0.73). In blood, however, colocalization provided little support for shared causal variant (ADHD-expression: H□ = 0.001; migraine-expression: H□< 0.001) (Table S14). Figure 4 shows the regional association patterns for MANBA expression and the corresponding ADHD and migraine GWAS signals at the MANBA locus.

**Figure 4.**
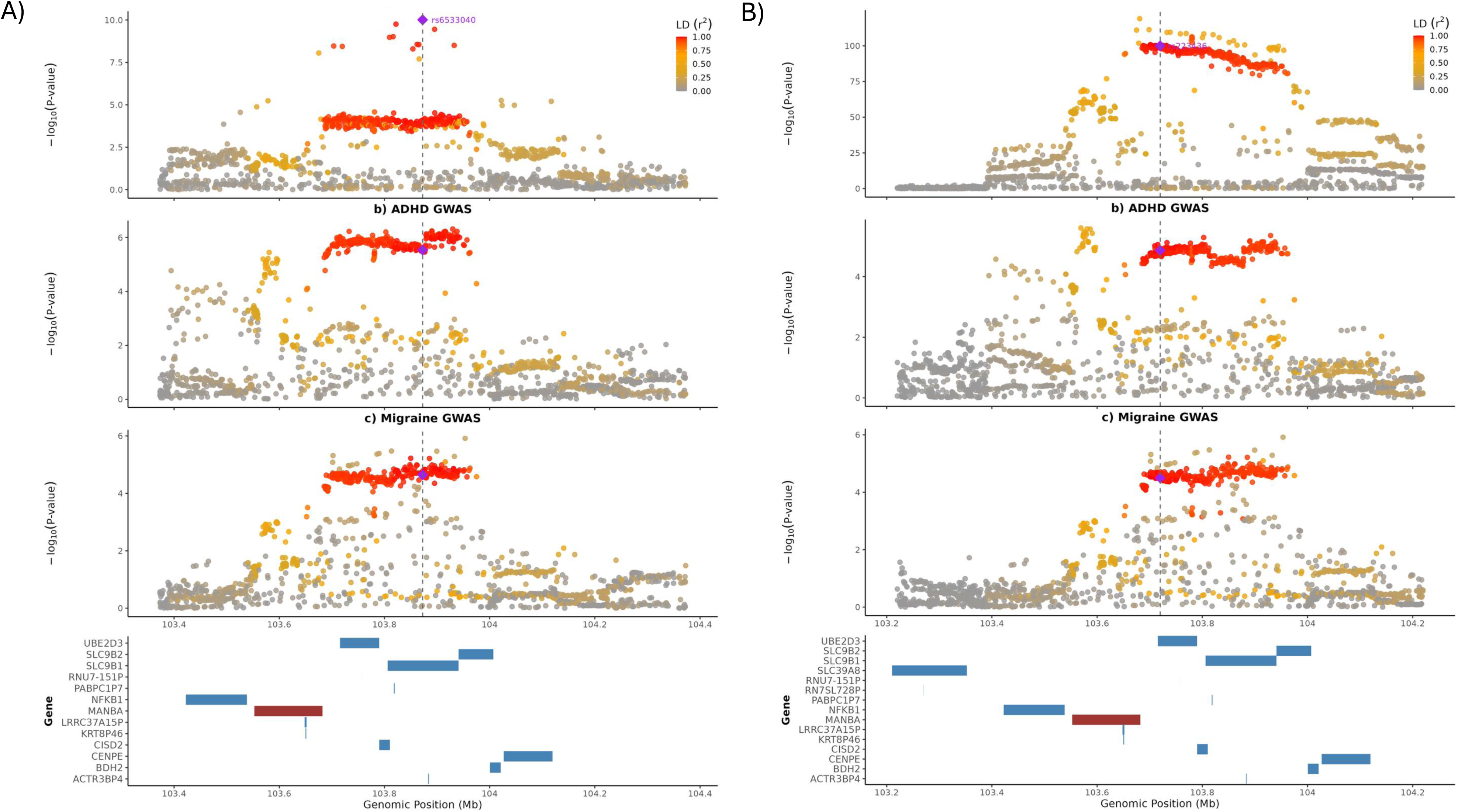
Regional association plots for MANBA expression in cortex (A) and blood (B) alongside ADHD and migraine association signals. SNPs are coloured according to linkage disequilibrium (LD, r²) with the lead eQTL variant.

## Discussion

In this study, we applied different approaches to assess the shared genetic background and molecular mechanisms underlying ADHD and migraine. Variant-level analyses identified multiple pleiotropic variants shared between the two conditions, including 20 variants with concordant effect directions. Regional analysis further identified a shared locus on chromosome 11. Cis-eQTL MR analyses of druggable genes implicated *MANBA* on chromosome 4 in both brain cortex and blood. Colocalization analyses supported a shared association signal in brain cortex, whereas blood showed evidence for distinct association signals for both ADHD and migraine.

At the variant level using conjFDR, we identified 27 loci jointly associated with ADHD and migraine, with 20 showing concordant effect directions. One pleiotropic locus tagged by rs56262138 exhibited opposite allelic effects, with colocalization analyses supporting discordant pleiotropic effects driven by a shared underlying signal. Gene-based analysis of this variant using MAGMA highlighted *CADM2* and *VGLL3* as candidate genes. Although neither gene was included in our MR analysis as they are not classified as druggable in DGIdb, previous genetic studies may still help contextualize the observed overlap between ADHD and migraine. *CADM2* has been repeatedly associated with impulsivity, risk-taking behaviour, substance use, and ADHD-related traits in large-scale GWASs [31]. The effect direction for this variant differs between ADHD and migraine, indicating that the shared signal at this locus does not imply concordant effects on both traits. This is consistent with PheWAS evidence showing trait-specific associations for CADM2-linked variants, including lower ADHD liability but higher migraine susceptibility [32]. Such opposing allelic effects may arise because shared genetic variants operate within different developmental and biological contexts across the lifespan. While ADHD reflects early-onset neurodevelopmental liability, migraine risk emerges later and is shaped by additional hormonal and environmental influences [33], potentially leading to divergent phenotypic effects of the same loci. Another gene located around this variant, *VGLL3*, is a transcriptional co-activator involved in inflammatory gene regulation [34]. Although its role in ADHD and migraine is less well characterised, increasing evidence suggests that immune and inflammatory processes may contribute to both conditions [35, 36]. Therefore, *VGLL3* may represent a potential link between inflammatory regulation and the shared genetic liability underlying ADHD and migraine.

Our local genetic correlation analysis using LAVA identified one genomic region on chromosome 11 (11: 112755447-113889019) showing a positive local genetic correlation between ADHD and migraine. Despite the absence of genome-wide significant variants, both ADHD and migraine showed significant local heritability in this region. This pattern suggested that the shared signal may arise from the cumulative effects of multiple subthreshold variants rather than a single pleiotropic variant [37], which may explain why this region was not consistently detected in analyses focused on identifying individual pleiotropic variants or strong colocalization signals. ADHD also demonstrated stronger association signals than migraine within this region. Previous LAVA analyses have similarly implicated this region across multiple neuropsychiatric and neurodegenerative traits despite limited evidence from global genetic correlation analyses, supporting the presence of shared local polygenic architecture within this locus [38].

When evaluating the causal effects of genetically predicted gene expression on ADHD and migraine, *MANBA* was the only gene supported by MR evidence for both traits. Mechanistically, *MANBA* encodes a lysosomal β-mannosidase central to glycoprotein turnover and autophagy [39]. Experimental data show that reduced *MANBA* activity disrupts lysosomal structure and impairs endocytosis and autophagy, whereas higher activity may enhance cellular clearance and attenuate inflammatory signalling [40], mechanisms that have been implicated in both ADHD and migraine [41, 42]. Consistent with this biology, prior transcriptomic and proteomic studies have linked *MANBA* to neuropsychiatric and neurological traits. At the transcriptomic level, previous S-PrediXcan study of ADHD and its comorbidities identified an association between *MANBA* expression in the cerebellar hemisphere and ADHD, whereas no association was observed for migraine [43]. At the protein level, MR leveraging pQTL for plasma β-mannosidase activity (the enzyme encoded by MANBA), reported a protective association with ADHD [44], with weaker but directionally consistent evidence observed for migraine [45]. In the present study, genetically predicted MANBA expression in cortex and blood showed associations with both traits, extending prior findings to a shared cross-trait context.

Colocalization analyses yielded tissue-specific results, with suggestive evidence for a shared causal variant in cortex but evidence favouring distinct causal variants in blood, indicating heterogeneity in the regulatory architecture linking *MANBA* expression to disease risk. In addition, a conjFDR-identified variant (rs6839635), although not meeting formal colocalization criteria (H4 = 0.69), was in high LD with the lead instruments for MANBA expression in both tissues, providing contextual variant-level support for involvement of the MANBA regulatory region. Taken together, these findings highlight MANBA as a biologically plausible candidate for further functional investigation [46, 47], while also suggesting a complex and potentially tissue-specific regulatory architecture underlying the observed associations.

This study has several limitations. First, the GWAS datasets included predominantly individuals of European ancestry, which may limit the generalizability of our findings to other populations. Second, differences in statistical power across analytic approaches may partly explain the limited convergence observed between findings. ConjFDR was designed to detect shared cross-trait enrichment and may be more sensitive to shared subthreshold associations, whereas follow-up PWCOCO relies more heavily on strong regional association signals and may therefore have been less sensitive to weaker or more complex shared loci.

Third, tissue-specific MR analyses were restricted to cortex and blood because of data availability and sample size, and therefore associations in other brain regions were not evaluated. Finally, gene-level analyses were restricted to druggable genes, which increased the translational relevance and reduced multiple-testing burden by prioritizing genes with pharmacological potential. However, many biologically important genes for psychiatric and neurological traits may not be classified as druggable, and restricting analyses to this subset might have missed relevant mechanisms. Therefore, the gene-level findings should be interpreted as target-focused rather than exhaustive.

Our findings suggest that ADHD-migraine comorbidity is unlikely to be driven by a single shared variant or pathway. Instead, different analytic approaches highlighted distinct components of the shared architecture, including pleiotropic variants, a locally correlated region on chromosome 11, and cis-eQTL signals involving *MANBA*. The limited overlap between these findings may reflect the distributed and heterogeneous nature of shared genetic risk, where variant-level associations, regional correlation, and genetically predicted gene-expression regulation capture different biological features of the ADHD-migraine relationship. In particular, the regional signal identified by LAVA and the tissue-specific cis-eQTL for *MANBA* suggest that regulatory mechanisms may contribute to the overlap beyond individual shared variants alone. Together, these results supported a model in which ADHD-migraine comorbidity arises through multiple partially overlapping mechanisms rather than a single shared biological pathway.

## Data Availability Statement

All data analysed in this study are publicly available. Summary statistics for ADHD are available for download through GWAS Catalog (ebi.ac.uk/gwas/studies/GCST90568441). Migraine GWAS summary statistics are available to bona fide researchers from the corresponding investigators of the original study. Brain eQTL data were obtained from the MetaBrain consortium and are publicly available through the MetaBrain portal (https://www.metabrain.nl/cis-eqtls.html). Blood eQTL data were obtained from the eQTLGen Consortium and are publicly available through the eQTLGen website (https://www.eqtlgen.org/cis-eqtls.html). No new data were generated during this study.

## Acknowledgements

This publication is the work of the authors, who will serve as guarantors for the contents of this paper. YL, ES (MC_UU_00032/1), CD (MC_UU_00032/6) are supported by the Medical Research Council Integrative Epidemiology Unit at the University of Bristol. YL is also funded by the China Scholarship Council (No. 202206240022). REW is funded by a postdoctoral fellowship from the South-Eastern Norway Regional Health Authority (2020024). CD is also funded by a postdoctoral fellowship from the South-Eastern Norway Regional Health Authority (2024078). For the purpose of Open Access, the author has applied a CC BY public copyright licence to any Author Accepted Manuscript version arising from this submission.

## Author Contribution Statement

YL conceived the study, performed the analyses, interpreted the results, and drafted the manuscript. ES, CD and REW contributed to the study design and supervision, interpretation of findings, and critical revision of the manuscript. All authors reviewed, edited, and approved the final manuscript.

## Ethical Approval

No new ethical approval was required for this study because all analyses were conducted using publicly available, anonymised GWAS summary statistics. Ethical approval and informed consent had been obtained by the investigators of the original studies from which the summary statistics were derived.

