## Supplementary Note for "Genomic analysis identifies polygenic and region-specific contributions to ADHD-migraine comorbidity"

### **Data Source**

The genome-wide association study (GWAS) data for ADHD were obtained from the latest meta-analysis [1], which combined GWAS summary statistics of quantitative ADHD symptom scores from 28 population-based cohorts (70,953 individuals; 290,134 observations) with clinical ADHD diagnosis data from the Psychiatric Genomics Consortium (38,691 cases and 186,843 controls) [2]. This combined meta-analysis (ADHD_OVERALL_) represents the most comprehensive investigation of ADHD genetic architecture to date. Details on case ascertainment, genotyping, and quality control are available in the original publication.

The GWAS data for migraine were obtained from a large-scale meta-analysis of publicly available datasets, comprising 48,975 self-reported/International Classification of Headache Disorders second edition (ICHD-II) cases and 540,381 population controls [3]. This comprehensive analysis aggregated results from four major study collections: the International Headache Genetics Consortium (IHGC) 2016 dataset (consist of 21 independent cohorts), the UK Biobank (UKBB) cohort, the GeneRISK study, and the Trøndelag Health Study (HUNT). Detailed information regarding genotyping platforms, quality control procedures, and analytical methodologies is available in the original publication [3].

### **Supplementary Method**

#### S1. Conjunctional FDR, variants-level colocalization, and functional annotation

S1.1 Conjunctional False Discovery Rate

We applied conjunctional false discovery rate (conjFDR) to identify variants jointly enriched for association with ADHD and migraine. ConjFDR increases sensitivity for detecting cross-trait pleiotropic variants by leveraging the empirical enrichment of association signals across both GWAS datasets[4, 5]. For each SNP, we compute two conditional FDR values: the FDR for ADHD given its association ranking for migraine, and the FDR for migraine given its association ranking for ADHD. The conjFDR for a SNP is defined as the maximum of these two conditional FDRs, providing a conservative measure of joint significance and allowing detection of pleiotropic variants that may not reach genome-wide significance in either GWAS individually. The conjFDR is derived from the maximum P value, calculated as:

$conjFDR=max(condFDR_{ADHD|MIG},condFDR_{MIG|ADHD}$)

Importantly, conjFDR is robust to differences in statistical power and sample size between the two GWAS. Because the method relies on enrichment rather than absolute P-value thresholds, strong signals from the higher-powered trait can increase the probability that SNPs with modest associations in the lower-powered trait represent true shared genetic architecture. As demonstrated in the original methodological work, conjFDR maintains appropriate control of false discoveries while substantially improving sensitivity under asymmetric power conditions.

S1.2 Colocalization of Pleiotropic Variants (PP.H4 > 0.8)

To distinguish true cross-trait pleiotropic from linkage disequilibrium (LD) between distinct causal variants, we performed pairwise conditional colocalization (PWCOCO) on all variants identified through conjFDR. PWCOCO extends standard Bayesian colocalization by conditioning on secondary association signals in each region, thereby allowing evaluation of whether two traits share the same causal variant after accounting for multiple independent signals. This approach reduces false colocalization due to LD structure or multiple causal traits with the same locus and improves specificity for shared causal genetic architecture [6]. Posterior probability of a shared causal variants (PP.H4) was used as the primary result, with PP.H4>0.8 interpreted as strong evidence for colocalization.

S1.3 Gene-Level Context for Colocalized Variants

Colocalized variants were annotated using Multi-marker Analysis of GenoMic Annotation (MAGMA) [7]. For SNP-to-gene mapping, SNPs were assigned to genes based on physical boundaries using the exact gene body. MAGMA was then applied separately for ADHD and migraine to annotate the genes surrounding the pleiotropic variants. MAGMA aggregates SNP-level association statistics into gene-level tests by modelling the joint effects of variants within each gene while accounting for local LD, thereby providing gene-level context for the pleiotropic loci.

S1.4 Interpretation of Variant-Level Results

The combined conjFDR, pairwise conditional colocalization, and MAGMA analyses characterize the variant-level architecture underlying the shared genetics between ADHD and migraine. Variants identified through conjFDR represent loci with cross-trait enrichment, consistent with polygenic pleiotropy rather than isolated genome-wide significant SNPs. Conditional colocalization further confirmed that a subset of these pleiotropic variants reflects shared causal signals rather than linkage disequilibrium between distinct association peaks. Gene-based aggregation using MAGMA provided additional context for these colocalized loci by summarizing the cumulative SNP-level signal at the gene level.

#### S2. Local Genetic Architecture Analyses Using LAVA

S2.1 Local cross-trait genetic correlation across LD-defined genomic regions.

LAVA was used to quantify local SNP-based heritability and shared genetic correlation between ADHD and migraine. The analysis proceeded in two stages: (1) estimation of univariate local heritability for ADHD and migraine, followed by (2) bivariate local correlation analysis in regions where both traits exhibited significant local heritability. Because the statistic reflects covariance rather than marginal significance, LAVA can detect shared regional architecture even when one trait shows stronger association signals than the other, making it particularly sensitive to concordant effect-size patterns.

Univariate local heritability (h² local) was first estimated for each trait, and loci showing evidence of local heritability in both ADHD and migraine (P<0.05/2495) were retained for bivariate analysis. We then calculated local genetic correlations (ρ-local) between the two traits to characterize shared regional genetic architecture. The significance of the bivariate correlations was corrected for multiple testing using the FDR procedure. UK Biobank European sample was used as reference panel, which provides a large-sample LD structure optimized for European-ancestry data and reduces bias compared to the 1000 Genomes reference [8].

S2.2 Gene-Based Analysis Within the LAVA-Defined Region

To provide gene-level aggregation of association signals within the region identified by LAVA, we performed a region-restricted gene-based analysis using MAGMA. The analysis window was defined strictly by the boundaries of the LAVA LD block that showed significant local genetic correlation between ADHD and migraine. All SNPs falling within this LAVA-defined region were assigned to genes based solely on genomic overlap with the region, and MAGMA gene-based association statistics were computed separately for ADHD and migraine. This approach provides a gene-level summary of association signals within the LAVA-defined region for each trait.

S2.3 Interpretation of Regional-Level Results

The LAVA analysis identified genomic regions where ADHD and migraine share local genetic architecture. Unlike variant-level approaches, LAVA quantifies aggregated regional heritability and does not require individual SNPs or genes within the region to exhibit genome-wide significance. A significant local genetic correlation therefore indicates that the joint genetic signal within this region contributes to both traits. This pattern is consistent with locus-level polygenic sharing, in which multiple variants collectively contribute to the regional signal rather than a single dominant causal variant.

#### S3. Druggable Mendelian Randomization and Tissue-Specific Colocalization

S3.1 Identification of druggable genes

Druggable genes were obtained from the Drug-Gene Interaction Database (DGIdb V.5.0.10, <https://www.dgidb.org/>). The DGIdb provides information on drug-gene interactions and druggable genes from publications, databases and other web-based sources. We downloaded the ‘Categories Data’ (released in December 2024), including all genes in the druggable categories in the DGIdb, from all sources mapped to Entrez genes [9]. Genes annotated within DGIdb’s druggable categories were included as the candidate set for MR analysis.

S3.1 Druggable Gene MR Using Tissue-Specific eQTLs

We performed druggable Mendelian Randomization using cortex and blood eQTL datasets. Gene expression served as the exposure, with eQTL variants as instruments. Conducting MR in two tissues allows evaluation of central nervous system-specific effects (cortex) as well as blood-based regulatory mechanisms that may contribute to ADHD and migraine.

S3.2 Colocalization of MR-Supported Genes

To evaluate whether the MR-identified associations between gene expression and ADHD or migraine reflected shared causal variants, we performed pairwise conditional colocalization only for the tissue-trait combinations that showed significant MR effects. For each MR association, the colocalization locus was defined as a ±500 kb window around the corresponding lead eQTL instrument used in the MR analysis. Colocalization was then conducted between the tissue-specific eQTL data (cortex or blood) and the GWAS summary statistics for the relevant trait (ADHD or migraine). Conditional colocalization models were applied to account for multiple independent signals within each locus. Posterior probability for a shared causal variant (PP.H4) was used as the primary statistic, and PP.H4 > 0.8 was considered strong evidence supporting a shared causal variant underlying both the eQTL and trait association signals.

S3.3 Interpretation of Druggable MR and Tissue-Specific Colocalization Results

The druggable MR analyses identified genes whose genetically predicted expression in cortex or blood is associated with ADHD or migraine. Colocalization applied to the corresponding tissue-trait pairs helped distinguish associations driven by shared causal variants from those potentially influenced by LD. Genes supported by both MR and colocalization therefore represent more reliable candidates whose regulatory variation may contribute to disease risk. These tissue-specific, genetically anchored targets complement the variant- and region-level findings by providing gene-level hypotheses with potential biological and therapeutic relevance.

### **Supplementary Figure**

#### Figure S1. Conditional Q-Q plot showing cross-trait enrichment between ADHD and migraine.

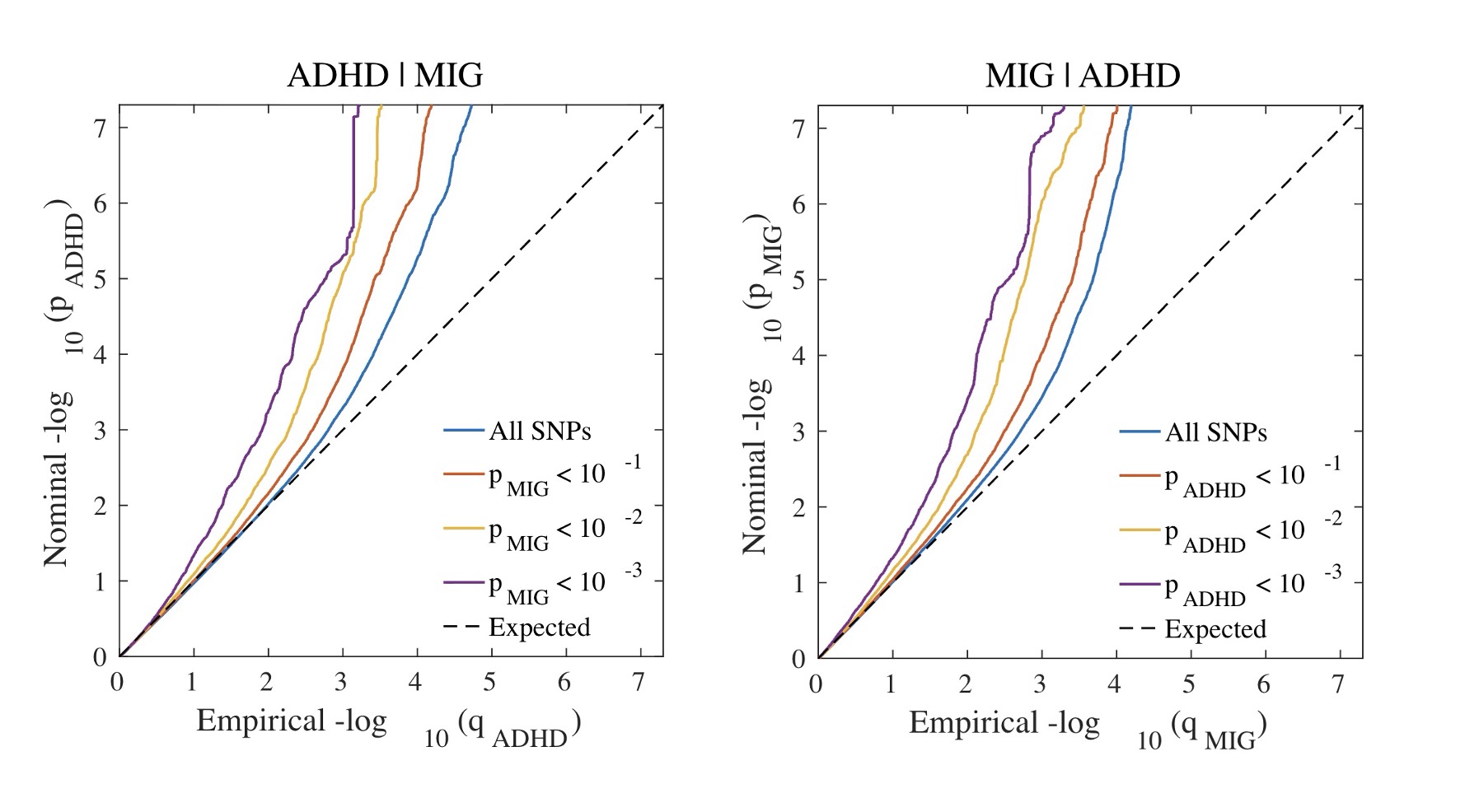

Figure S2. Pairwise scatter plot for ADHD and migraine around SNP rs56262138.

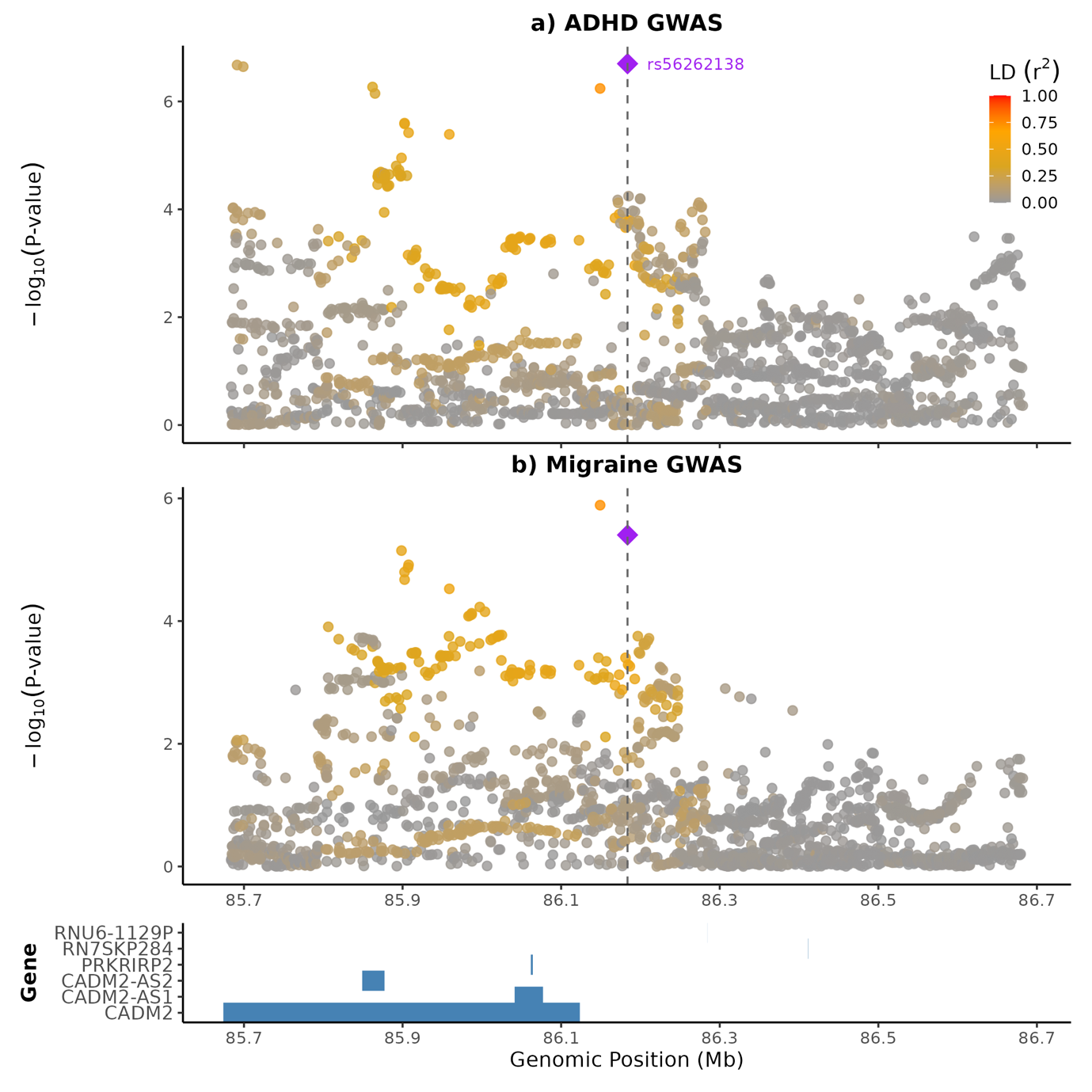

#### Figure S3. Pairwise scatter plot for ADHD and migraine around SNP rs2431108.

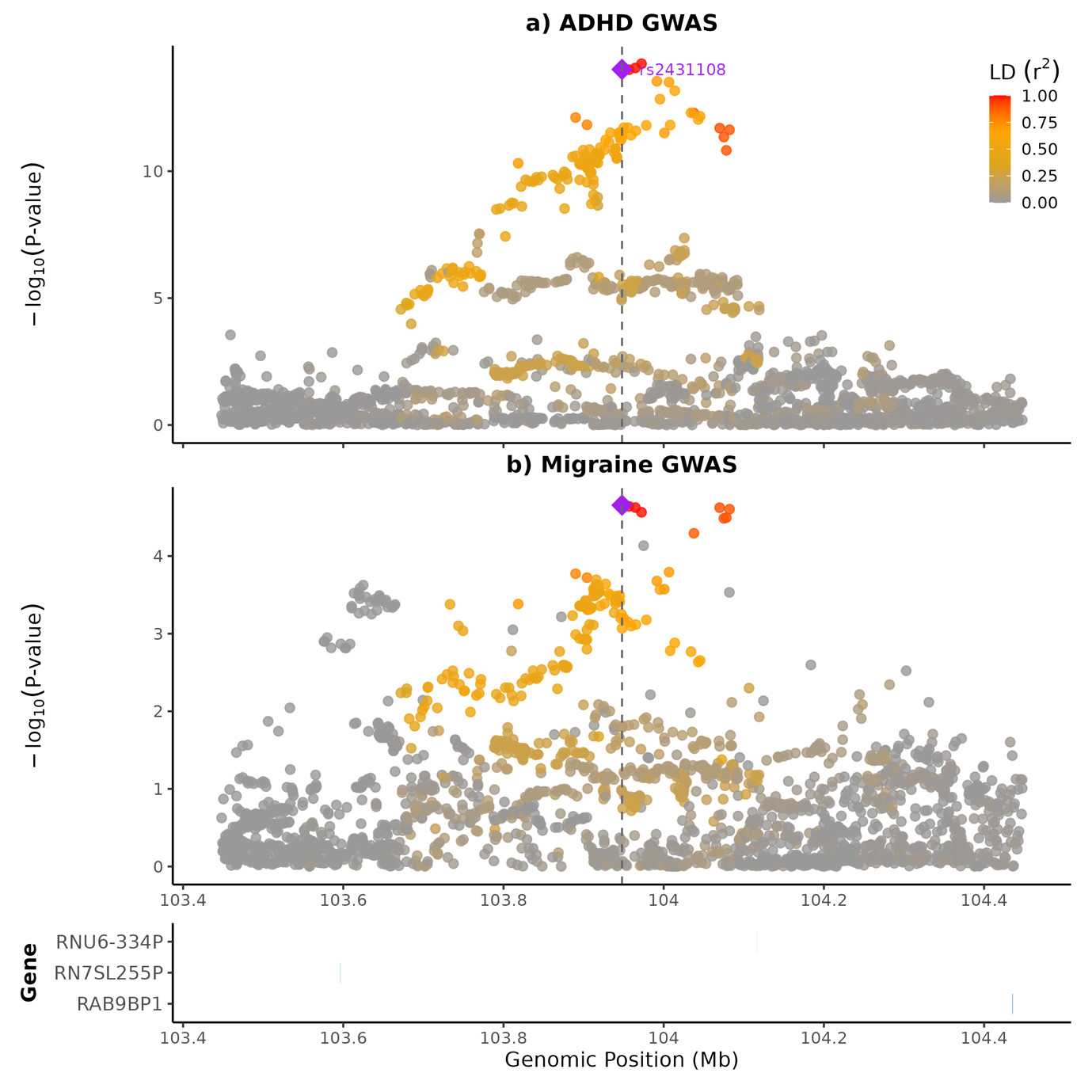
